# USA Winter 2021 CoVID-19 Resurgence Post-Christmas Update

**DOI:** 10.1101/2022.02.04.22270491

**Authors:** Genghmun Eng

## Abstract

We have successfully modeled every USA CoVID-19 wave using:

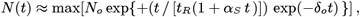

where *N* (*t*) is the total number of new CoVID-19 cases above a prior baseline, and *t*_*R*_ sets the doubling time *t*_*dbl*_ = *t*_*R*_ (ln 2). The new parameters {*α*_*S*_ ; *δ*_*o*_} measure mitigation efforts among the *uninfected population*, with {*α*_*S*_ > 0} being associated with *Social Distancing* and *vaccinations* ; while {*δ*_*o*_ > 0 }is associated with *mask-wearing*, which gives a faster 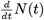 post-peak *drop-off*. The predicted pandemic wave end is when *N* (*t*) no longer increases.

Using data from 11/15/21-12/30/21, our prior *medrxiv*.*org* preprint* showed this initial *Omicron* CoVID-19 wave had values that matched the *initial* stage of the prior USA Winter 2020 resurgence {*t*_*R*_ ≈8 05 *days* ; *α*_*S*_ ≈ 0 011 */ day*}, when practically no one was vaccinated. In addition, this *initial* Winter 2021 wave showed virtually no *mask-wearing* {*δ*_*o*_ *≈ ×* 0 001 10^−3^ / *day*}, making it capable of infecting virtually everyone. These parameter values indicated that the *Omicron* variant was likely evading the vaccines in people who thought they were protected.

As a result, stopping the *Omicron* CoVID-19 spread must once again rely on enhanced *Social Distancing* and *mask-wearing*, just like the initial pandemic wave in March 2020. Analyzing the USA follow-on data from 12/25/21-1/31/22 shows that people did exactly that after the *Christmas Holiday* season, resulting in the following model parameters and values:

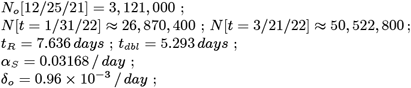

for this wave by itself, with all prior waves subtracted out as a baseline. Combining all the USA CoVID-19 waves gives these updated totals:

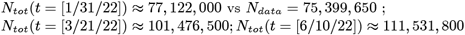

assuming no future CoVID-19 Resurgence (*with 4 Figures*).

*\*(10*.*1101_2021*.*10*.*15*.*21265078)*

## 1 Introduction

Each USA CoVID-19 wave**^1-7^**, from the pandemic start (3/21/20) to the present day (1/31/22), has been successfully modeled using this basic *N* (*t*) function for the total number of new CoVID-19 cases above a prior baseline:

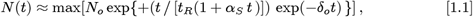

with *t*_*R*_ setting the pandemic doubling time *t*_*dbl*_ = *t*_*R*_ (ln 2), as in standard **SEIR** (**S**usceptible, **E**xposed, **I**nfected, **R**ecovered or Removed) epidemiology models. The new Eq. [1.1] parameters model mitigation efforts among the *uninfected population*, with {*α*_*s*_ > 0} associated with *Social Distancing* and *vaccinations*; and {*δ*_*o*_ > 0} associated with *mask-wearing*, which gives a faster 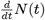 post-peak *drop-off*. The predicted pandemic wave ends when *N* (*t*) no longer increases.

Given a total population of *N*_*ALL*_, the *uninfected population U* (*t*) is:

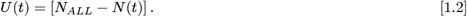

Using Eq. [1.1] assumes *N* (*t*) << *N*_*ALL*_, so that pandemic saturation effects can be ignored. Also, **SEIR** models do not generally include what the *U* (*t*) *uninfected population* is doing in response to the pandemic. In contrast, Eq. [1.1] was developed as a *non-local* extension of **SEIR** models, to account for how the *uninfected population*, as a whole, is mitigating the pandemic spread.

Each new USA CoVID-19 wave starts with a sharp rise in the total number *N* (*t*) of new cases, while the *t* = 0 point is chosen to be when the resurgence is first easily identified, with *N* (*t* = 0) = *N*_*o*_ being the number of cases above baseline at that time. Since Eq. [1.1] is empirically based, it does not predict when each new CoVID-19 wave will start, or what biological and social circumstances are causing the new wave. But once the CoVID-19 wave becomes established, Eq. [1.1] appears to successfully predict its time evolution.

Since the same few parameters in Eq. [1.1] have successfully modeled the time evolution of each USA CoVID-19 wave**^1-7^**, this result shows that the **response** of the *U* (*t*) *uninfected population* has been similar for each wave, even if different dominating factors were driving the CoVID-19 resurgences.

All CoVID-19 data used here came from the open-source *bing*.*com* CoVIDTracker**^8^** database. The initial stage of the USA Winter 2021 CoVID-19 Resurgence started when the *Omicron CoVID-19* variant became ascendant. Our prior *medrxiv*.*org* preprint**^7^** covering data from 11/15/21-12/30/21 showed that this stage was characterized by:

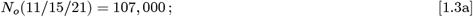

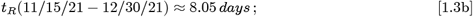

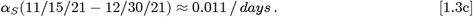

These values are similar to the USA Winter 2020 CoVID-19 Resurgence, which only had a fully-vaccinated rate of ∼0 3%. Since the USA now has a significantly vaccinated population, it makes it likely that the *Omicron CoVID- 19* variant is evading the vaccines.

While the prior 2020 Winter Resurgence had a significant amount of *maskwearing* [δ_*o*_(2020) 1 748 ×10^−3^ */ day*], the *initial* part of the 2021 Winter Resurgence was associated with virtually no *mask-wearing* :

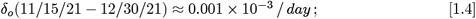

indicating that the *Omicron* variant was infecting people who thought they were protected. While *Omicron* vaccine evasion is now generally accepted, vaccination still offers protection against hospitalization from an *Omicron CoVID-19* infection. However, with virtually no *mask-wearing*, the Eqs. [1.3a]-[1.3c] values showed that this *initial* stage of the Winter 2021 CoVID-19 wave showed that it could infect virtually all susceptible people**^7^**.

This result similar to the *initial* stage of the USA Summer 2021 wave**^5^**. That commonality shows that resurgences may be driven by people letting their guard down with respect to *Social Distancing* and *mask-wearing*. As this *initial* Winter 2021 CoVID-19 Resurgence occurred between *Thanksgiving* and *Christmas*, festivities likely contributed to this CoVID-19 surge.

However, right after the *Christmas Holiday*, when the number of new cases reached ∼3, 120, 000 cases above baseline, the *uninfected population* began practicing enhanced *Social Distancing* and *mask-wearing*. This change is similar to what occurred during the latter portion of the USA Summer 2021 CoVID-19 wave**^5^**, and it is modeled here in this update.

## 2 Post-*Christmas* USA Winter 2021 Resurgence

Using the USA Summer 2021 CoVID-19 Resurgence as a baseline, it showed that the USA Winter 2021 CoVID-19 Resurgence started around 11/15/21. The dots in *Figure 1* show the total number of new CoVID-19 cases above the Summer 2021 USA CoVID-19 baseline**^5-6^**. The *Figure 1* continuous line shows the original CoVID-19 projections for this *initial* stage of the USA Winter 2021 CoVID-19 Resurgence**^7^**, using data from 11/15/21-12/30/21.

**Fig. 1:**
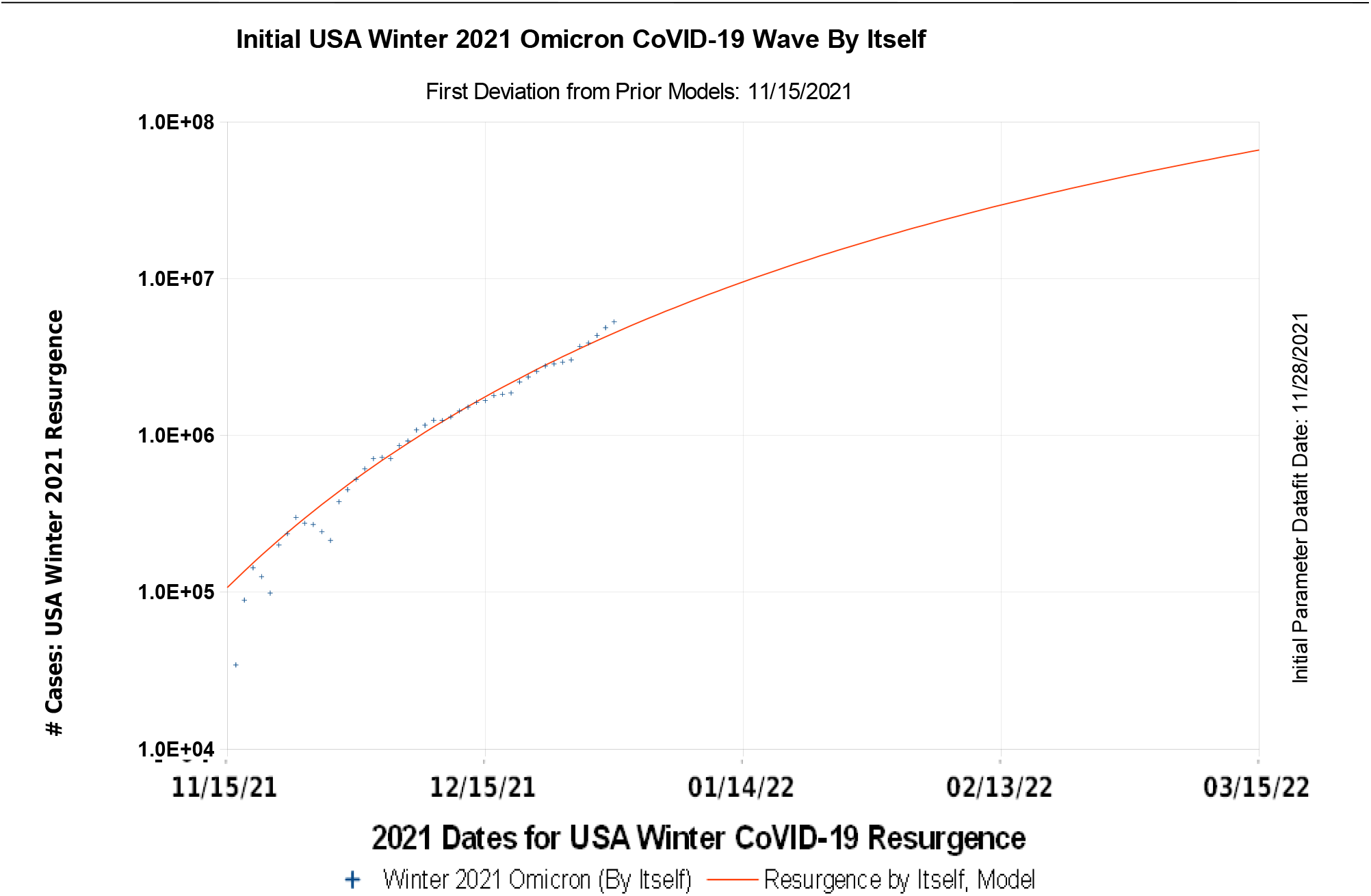
The Initial USA CoVID-19 Winter 2021 Resurgence, by Itself

*Figure 2* includes follow-on data after this *initial* USA Winter 2021 CoVID- 19 Resurgence, with data and models updates for 12/25/21-1/31/22, covering the post-*Christmas* CoVID-19 surge. Almost immediately after the *Christmas Holiday*, the *uninfected population* returned to enhanced *Social Distancing* and *mask-wearing*, creating a distinct change in the *N* (*t*) function after 12/25/21.

**Fig. 2:**
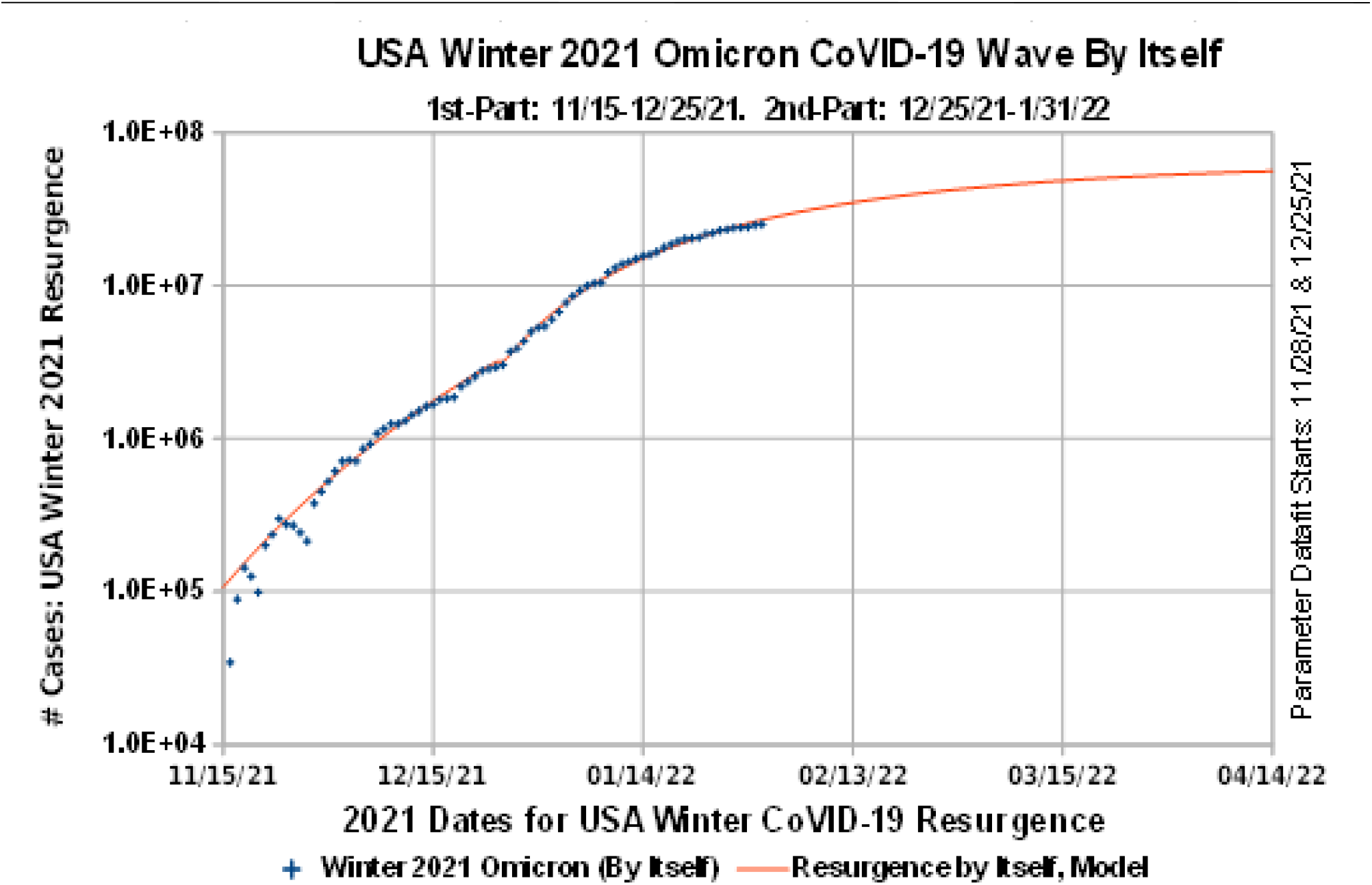
USA CoVID-19 Winter 2021 Resurgence Post-Christmas Update

Modeling the 12/25/21-1/31/22 period of the USA Winter 2021 wave in *Figure 2* shows that this CoVID-19 wave can subside prior to infection of the entire USA population. This *latter* portion has these model parameters:

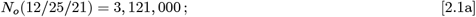

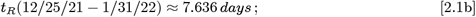

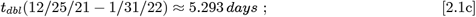

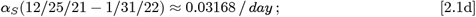

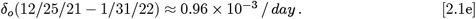

The *N*_*o*_(12/25/21) = 3, 121, 000 starting point for the behavior change in *N* (*t*) is similar to the prior Summer 2021 Resurgence, which also showed a behavior change in *N* (*t*) at *N*_*o*_(8/13/21) = 3, 200, 000. This commonality shows that the *uninfected population* altered their behavior at similar points, which is likely when some hospitals became overwhelmed.

*Figure 3* shows the total number of CoVID-19 cases, *N*_*tot*_(*t*), resulting from combining all waves of the CoVID-19 pandemic, from March 2020 through January 2022. Whenever the pandemic appeared to be beaten down, people became more complacent, allowing new CoVID-19 variants to spread, and the pandemic rose up again, almost with every season.

**Fig. 3:**
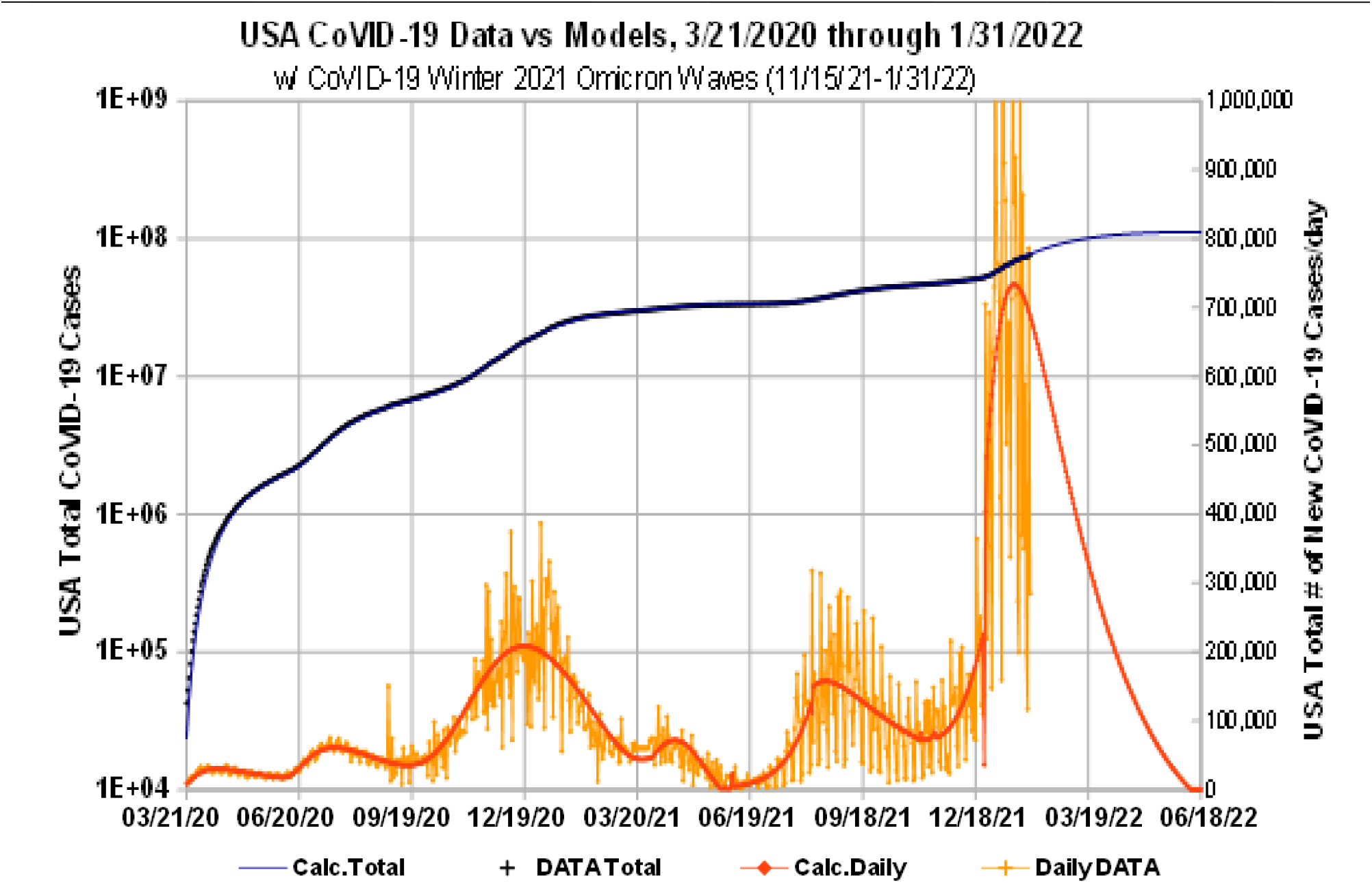
USA CoVID-19 Totals: 3/21/2020 through 1/31/2022

The daily number of new cases *dN*_*tot*_(*t*) */ dt* in *Figure 3* has peaks for the initial Spring 2020 pandemic; a Summer 2020 resurgence; the long Winter 2020 Resurgence; a small uptick in Spring 2021; the Summer 2021 Resurgence; and now the Winter 2021 Resurgence.

*Figure 4* presents a tabulated summary of all the model parameters that were derived for each CoVID-19 wave. It shows these overall features:

**Fig. 4:**
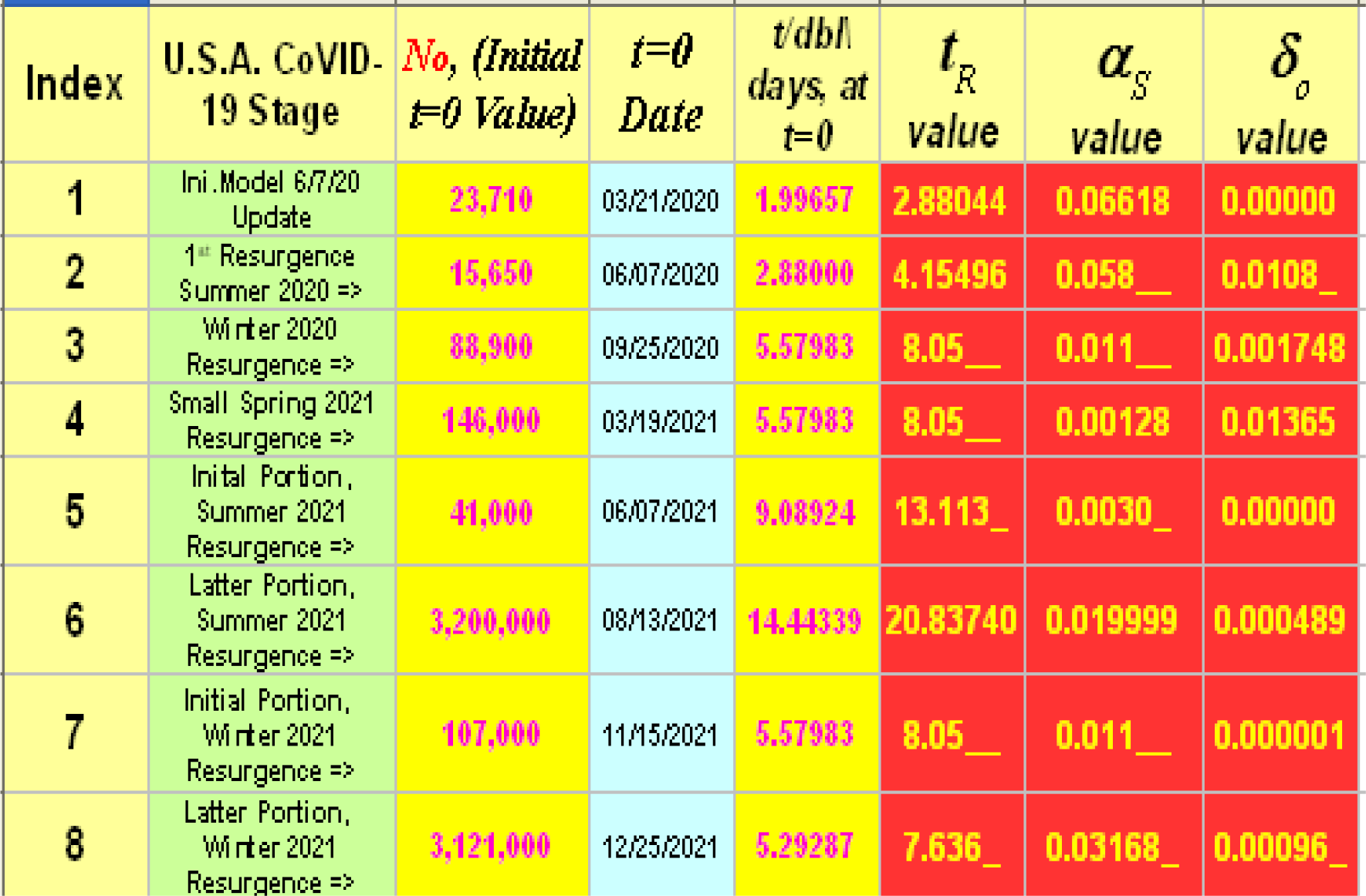
Summary of CoVID-19 Model and Parameter Values

a) The *t*_*dbl*_ = *t*_*R*_ (ln 2) doubling times for several CoVID-19 waves vary only from {5 3 - 5 6 *days*}.
b) The *a*_*s*_ *Social Distancing* and *vaccination* parameter for this CoVID-19 wave is one of the largest values, except for the early 2020 pandemic start.
c) Given a fast-rising CoVID-19 pandemic Resurgence, the *uninfected population* generally takes notice of the severity of a new CoVID-19 wave when the new infections reach a level of ∼3, 000, 000 above its baseline.
d) The *mask-wearing* parameter, *o*_*o*_, models the width and duration of the post-peak tail. Increased *mask-wearing* remains one the most powerful factors for hastening the CoVID-19 pandemic end.

## 3. Summary

Since our CoVID-19 modeling has been successful at predicting the time evolution of each USA CoVID-19 wave**^1-7^**, using the same few parameters:

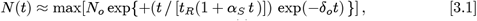

it indicates that the **response** of the *U*(*t*) uninfected population was similar for each wave, even if different dominating factors drive each new resurgence.

The initial stage of the USA Winter 2021 CoVID-19 Resurgence started with the *Omicron CoVID-19* variant becoming dominant. Our prior *medrxiv*.*org* preprint**^7^**, covering data from 11/15/21-12/30/21, showed that this *initial* stage was characterized by:

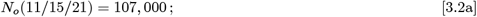

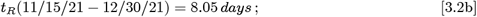

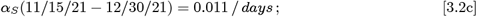

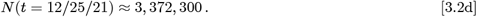

Thes {*N*_*o*_ *t*_*R*_; *α*_*s*_} _11/15/21_ arameters are very similar to the prior USA Winter 2020 CoVID-19 Resurgence, when the USA only had a fullyvaccinated rate of ∼0 3%. Since the USA now has a significantly vaccinated population, this new Winter 2021 Resurgence *Omicron CoVID-19* variant is likely evading the vaccines. Since the *N*_*o*_(12/25/21) starting point of Eq. [2.1a] was nearly six weeks after the Eq. [3.2a] starting point, those two values should be similar, as they are.

Just after the *Christmas Holiday*, the *uninfected population* responded to this CoVID-19 surge with enhanced *Social Distancing* and *mask-wearing*. The 12/25/21-1/31/22 post-*Christmas* data gives Eq. [2.1a]-[2.1e] as model parameters for this *latter* portion of the USA Winter 2021 CoVID-19 Resurgence, along with these updated *N* (*t*) predictions for this CoVID-19 wave by itself:

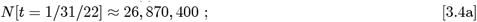

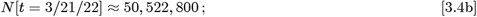

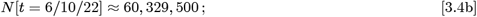

with this wave no longer infecting practically everyone. Combining these results with the prior *medrxiv*.*org* preprint**^7^** data, gives these updated USA CoVID-19 projections for the total number of USA CoVID-19 cases:

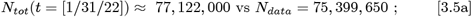

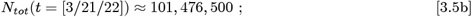

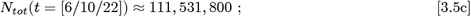

from all USA CoVID-19 waves, assuming no new CoVID-19 resurgence intervenes. More *mask-wearing* would further mitigate CoVID-19 spread, and it could significantly reduce these projected future USA CoVID-19 totals.

## Data Availability

All data produced in the present work are contained in the manuscript.

